# Impact of the COVID-19 pandemic on regional and national uptake of HIV testing services in Sierra Leone: a descriptive analysis

**DOI:** 10.1101/2025.01.21.25320892

**Authors:** Hannah Sheriff, Ginika Egesimba, Bailah Molleh, Jarieu Tucker, Gerald Younge, Francis K. Tamba, Sulaiman Lakoh, Adrienne K. Chan, Sharmistha Mishra, Stephen Sevalie

## Abstract

**Background:** A key milestone in the reduction of the global HIV burden is reaching the UNAIDS 95-95-95 target by 2025. The COVID-19 pandemic may have affected the ability of countries to achieve this target, but data to describe this impact is limited. This study assessed national HIV testing service uptake in Sierra Leone during three periods of the COVID-19 pandemic.

**Methods:** We conducted a retrospective cross-sectional study using secondary program data of all patients tested for HIV in all 16 districts of Sierra Leone. Data from March 2019 to February 2020 (pre-COVID-19); March 2020 to February 2021 (during COVID-19); and March 2021 to February 2022 (post-COVID-19) were extracted from DHIS-2 and descriptive analyses were performed using Stata (15.1, StataCorp LLC, College Station, TX).

**Results:** The median number of HIV tests was 58,588 (IQR 54,232 to 62,077) in the pre-COVID phase, 55,141 (IQR 52,975 to 57,689) during the COVID-19 phase, and 74,954 (IQR 72,166 to 76,250) in the post-COVID phase. This shows that HIV testing rate decreased by 6.3% during the COVID-19 period and increased substantially by 36.0% in the post-COVID-19 period. Twice more women than men were tested for HIV across all periods—pre-COVID-19 (38,825 vs. 19,789), during COVID-19 (36,923 vs. 17,755), and post-COVID-19 (49,205 vs. 25,472) phases.

**Conclusion:** Our study shows that HIV testing was significantly disrupted during the COVID-19 pandemic but recovered quickly after the pandemic. These findings highlight that lessons learned from previous epidemics may influence adaptive strategies to maintain essential health services during public health emergencies.

## Introduction

A key milestone in the reduction of the global burden of HIV is reaching the UNAIDS 95-95-95 target by 2025; 95% of all people living with HIV know their status, 95% of all people diagnosed with HIV receive sustained antiretroviral therapy and 95% of all people receiving antiretroviral therapy have viral suppression [1]. This global goal of controlling the epidemic has been severely impacted by the COVID-19 pandemic, particularly in low- and middle-income countries [2]. There are many factors that affect health service delivery during the pandemic. Lockdown and travel restrictions, supply chain disruptions, changes in health-care-seeking behaviors, reductions or closures of facility services, redeployment of health-care staff and limited community engagement have all led to disruptions in essential services [3, 4].

HIV testing remains the main entry point to HIV care and treatment, and its interruption can jeopardize the entire care continuum [ 5]. Data from 502 health facilities in 32 countries in Africa and Asia showed a significant decrease in HIV testing services (41%) and HIV referrals (37%) due to the COVID-19 pandemic [2].

Sierra Leone has a population of 7.5 million, and an estimated 77,273 people living with HIV, a prevalence of 1.73% [6]. Key populations, such as female sex workers, have the highest HIV infection rates [6,7]. There are geographic disparities in HIV infection rates in Sierra Leone, with higher rates in urban areas than in rural areas [7].

The COVID-19 pandemic may have affected the country’s progress towards epidemic control in 2025. As of December 2023, only 83% of people living with HIV had been diagnosed [7]. Similar disruptions to HIV testing services were reported during the 2014/2016 Ebola outbreak, and these effects may have continued until the COVID-19 pandemic in March 2020 [8].

The National AIDS Control Program, in collaboration with partners, rapidly implemented a mitigation strategy to ensure continued access to HIV testing amid the challenges posed by the COVID-19 pandemic. This strategy included the formulation of guidelines for integrating HIV testing services in the centers providing COVID-19 services, scaling up HIV testing services in COVID-19 treatment facilities, implementing COVID-19 symptom screening as a prerequisite for accessing HIV services, and imposing a temporary suspension on community-based testing. Given the differences in health service delivery and human resources across regions, the impact of these adaptive measures to the utilization of essential health services is expected to vary from region to region. However, a comprehensive evaluation of the effectiveness of these interventions in mitigating the impact of COVID-19 on HIV testing services at national and sub-national levels in Sierra Leone has not yet been conducted.

Our study aimed to understand the geographic variation of the impact of COVID-19 on the HIV testing service uptake amongst persons aged 15-49 across the five given regions of Sierra Leone by comparing three-time periods: pre-COVID-19, during, and post-COVID-19.

## Methods

### Study design and setting

We conducted a retrospective cross-sectional study using aggregated secondary program data for all patients who were tested for HIV in public and private health facilities across all the 16 districts in Sierra Leone.

The 16 districts are aggregated and administered into five regions: Eastern (Kailahun, Kenema, and Kono Districts), Western Area (Urban and Rural Districts), Northern (Bombali, Falaba, Koinadugu, and Tonkolili Districts), Northwestern (Kambia, Karene, and Port Loko Districts) and, Southern (Bo, Bonthe, Moyamba, Pujehun Districts) regions. HIV testing services are provided in 960 primary, secondary and tertiary health facilities across the 16 districts by the National AIDS Control Program of the Ministry of Health. Key HIV testing approaches implemented by the program consists of HIV counseling and testing, provider-initiated testing and counseling, index case testing, community-based testing and HIV self-testing.

### Data sources and variables

Data for this study was extracted on October 2, 2024 from the DHIS-2, a national health management information systems database consisting of secondary de-identified, aggregated data reported monthly from the health facilities providing HIV testing services into a Microsoft Excel. Patient-level data collected daily during service provision was entered into standardized data collection tools aligned with nationally developed reportable data elements and indicators, with quality assurance provided by the district and national monitoring and evaluation systems. We extracted monthly data from March 2019 to February 2022 to capture seasonal variations in service utilization and reflect HIV testing services over three time periods: March 2019 to February 2020 (before the COVID-19 pandemic); March 2020 to February 2021 (during the COVID-19 pandemic); March 2021 to February 2022 (post-COVID-19 pandemic). Variables extracted included number of people tested per month, gender, region, and region.

### Data management and analysis

Data in Excel format was cleaned, coded, and validated prior to analysis. We visualized national and regional variations in patterns in HIV testing services uptake before, during, and post-COVID-19 pandemic. We reported the median and inter-quartile range in monthly HIV testing services uptake by time periods at the national level for the total population, and stratified by gender using Stata (15.1, StataCorp LLC, College Station, TX).

### Ethics

Ethical approval was obtained from the Sierra Leone Ethics and Scientific Review Committee of the Ministry of Health Board with approval number 016/05/2023. Routine data were abstracted under a waiver of informed consent. The authors did not have access to information that could identify individual participants during or after data collection.

## Results

### Summary of the uptake of HIV testing services

The median number of HIV tests was 58,588 (IQR 54,232 to 62,077) in the pre-COVID phase, 55,141 (IQR 52,975 to 57,689) during the Covid-19 phase, and 74,954 (IQR 72,166 to 76,250) in the post-COVID phase. This shows that HIV testing rates decreased by 6.3% during the COVID-19 period and increased substantially by 36.0% in the post-COVID-19 period. Gender differences in the uptake of HIV testing were reported in all three phases of testing. Across all periods — before COVID-19 (38,825 vs. 19,789), during COVID-19 (36,923 vs. 17,755) and after COVID (49,205 vs. 25,472), women consistently received about twice as many tests as men. Nevertheless, both genders have seen similar drops in HIV testing during the COVID-19 pandemic **(Table 1)**.

**Table 1:** Summary of the uptake of HIV testing services over three-time periods in Sierra Leone (March 2019 to February 2022)

| HTS Uptake per month | Pre-COVID<br>Mar 2019 to Feb 2020 | During COVID<br>Mar 2020 to Feb 2021 | Post-COVID<br>Mar 2021 to Feb 2022 |
| --- | --- | --- | --- |

|  | 12 months |  | 12 months |  | 12 months |  |
| --- | --- | --- | --- | --- | --- | --- |
|  | Median | (IQR) | Median | (IQR) | Median | (IQR) |
| Total | 58588 | (54232, 62077) | 55141 | (52975, 57689) | 74954 | (72166, 76250) |
| Male | 19,789 | (19204, 21659) | 17,755 | (17474, 19692) | 25,472 | (24522, 26044) |
| Female | 38,825 | (34883, 4041) | 36,923 | (35451, 38465) | 49,205 | (47626, 50514) |

### Variation in the number of HIV testing services across gender and different time periods of the COVID-19 pandemic

The use of HIV testing services increased steadily before the pandemic, but this trend suddenly dropped at the beginning of the pandemic. However, during the early part of the COVID-19 pandemic, there was a sharp increase in HIV testing services (**Figure 1**). HIV testing rates among women are approximately twice as high as among men (**Figure 2**).

**Figure 1:**
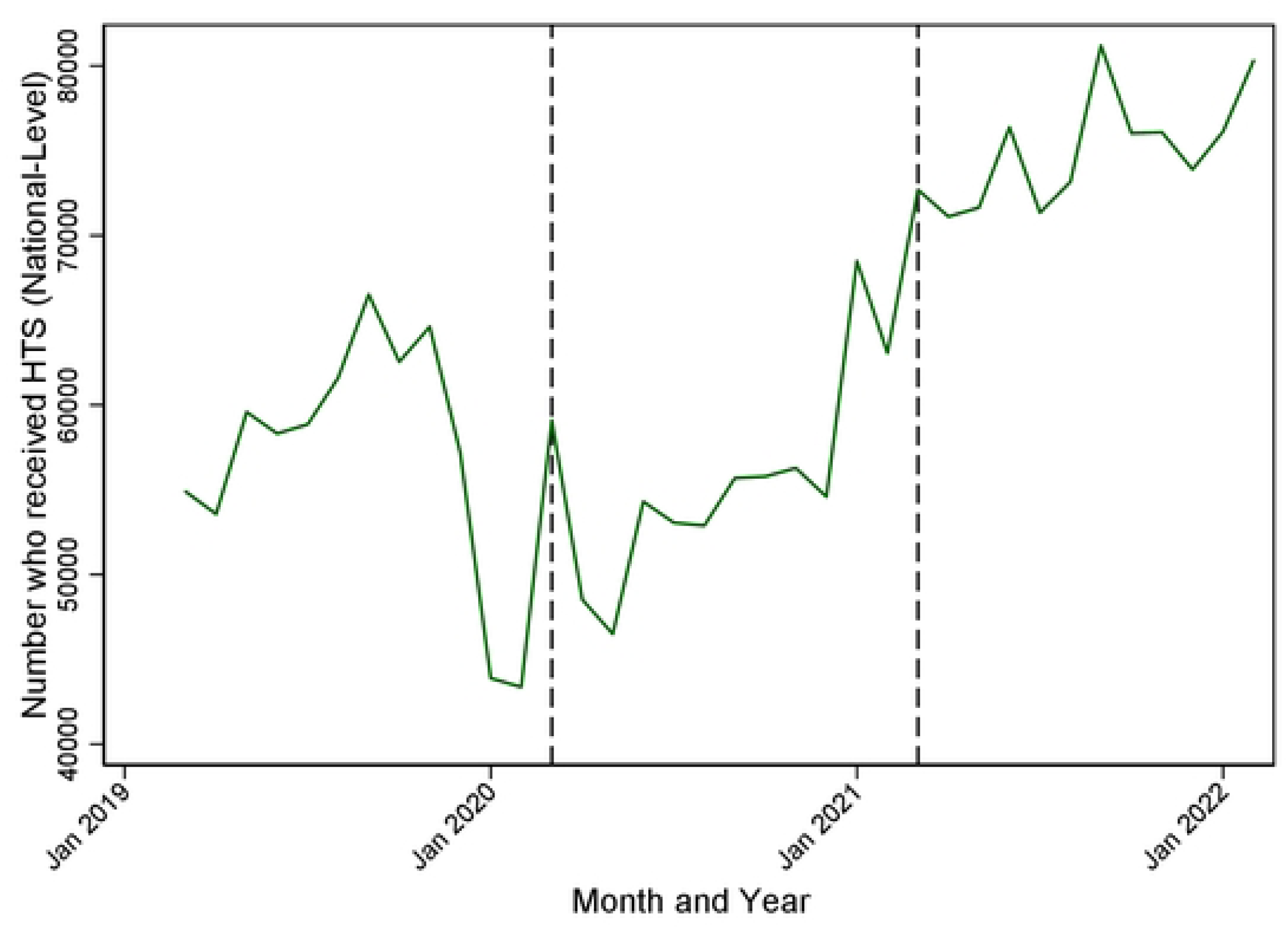
National HIV testing services uptake in three time periods

**Figure 2:**
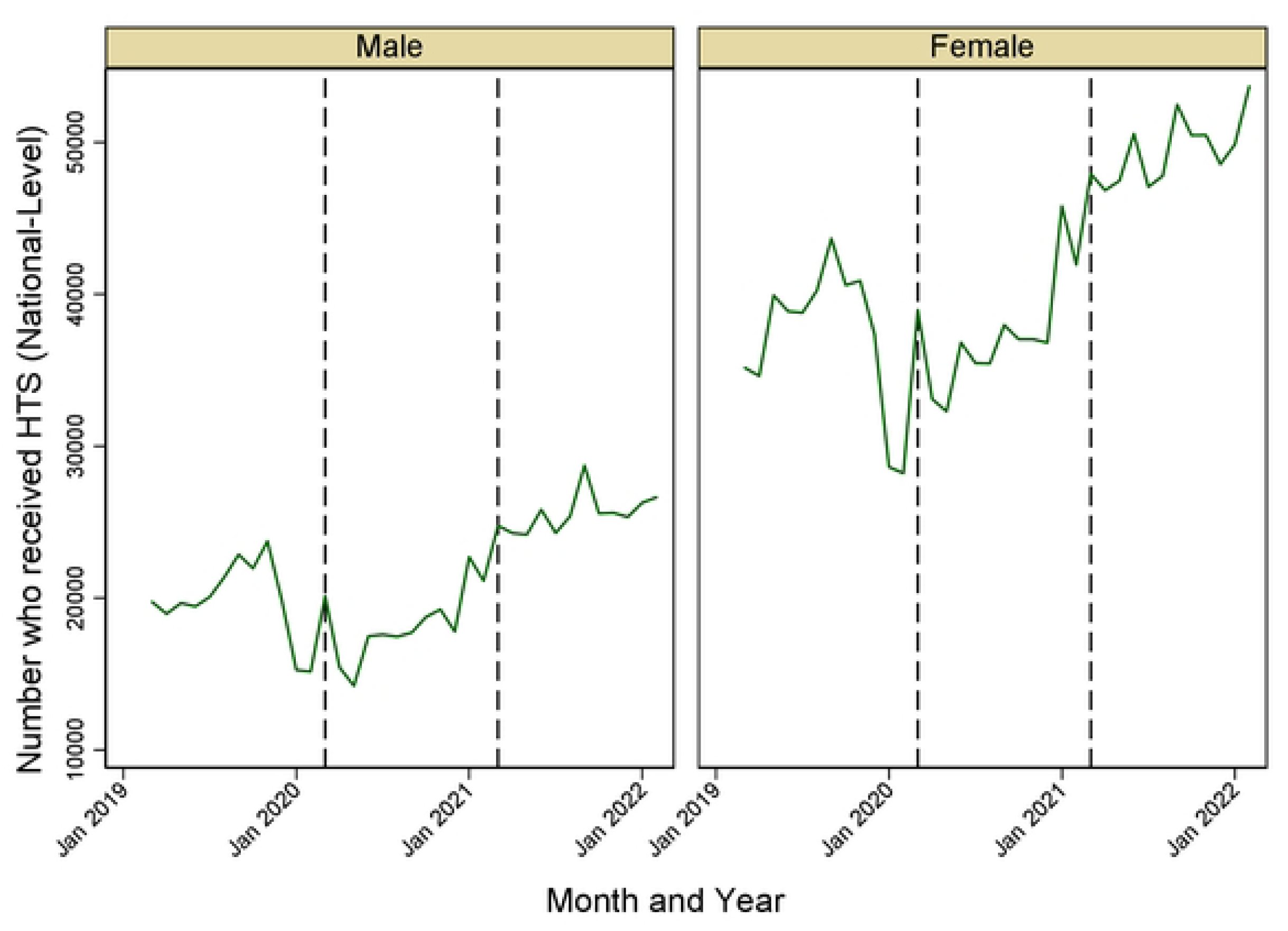
Variation in HIV testing during the COVID period by gender

### Variation in HIV testing services by region

There were minimal geospatial variations in the utilization of HIV testing services in the East, North, South, and Northwestern Regions across the different periods of COVID-19 pandemic. In contrast, the Western Area, characterized as the most urbanized part of Sierra Leone, exhibited a notable decrease in HIV testing just before the start of COVID-19 (**Figure 3**). The utilization of the HIV testing services in the Western Area Urban was highly variable, highlighting pronounced fluctuations attributed to COVID-19 pandemic (**Figure 4**).

**Figure 3:**
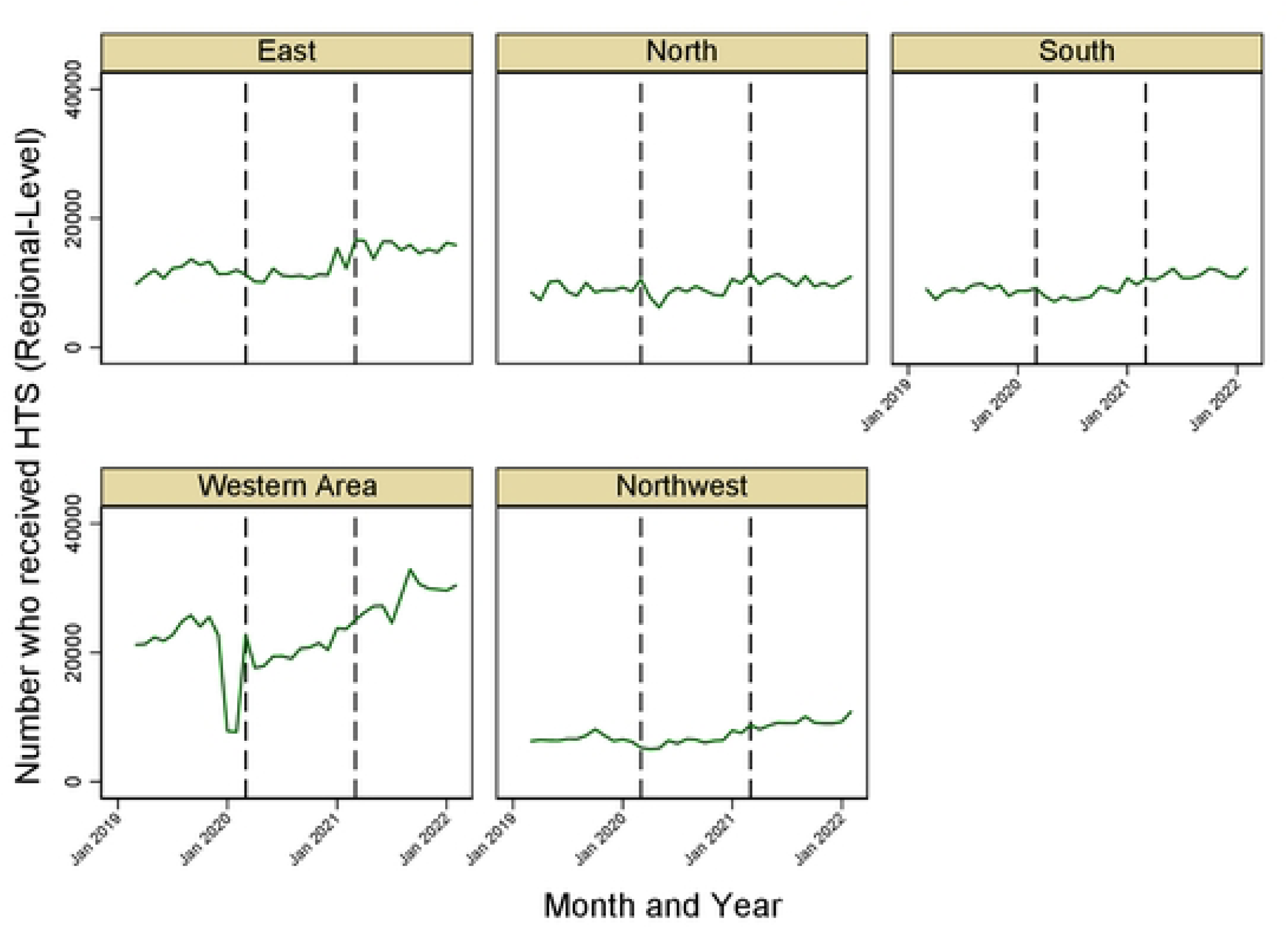
**Uptake of HIV testing services by region**

**Figure 4:**
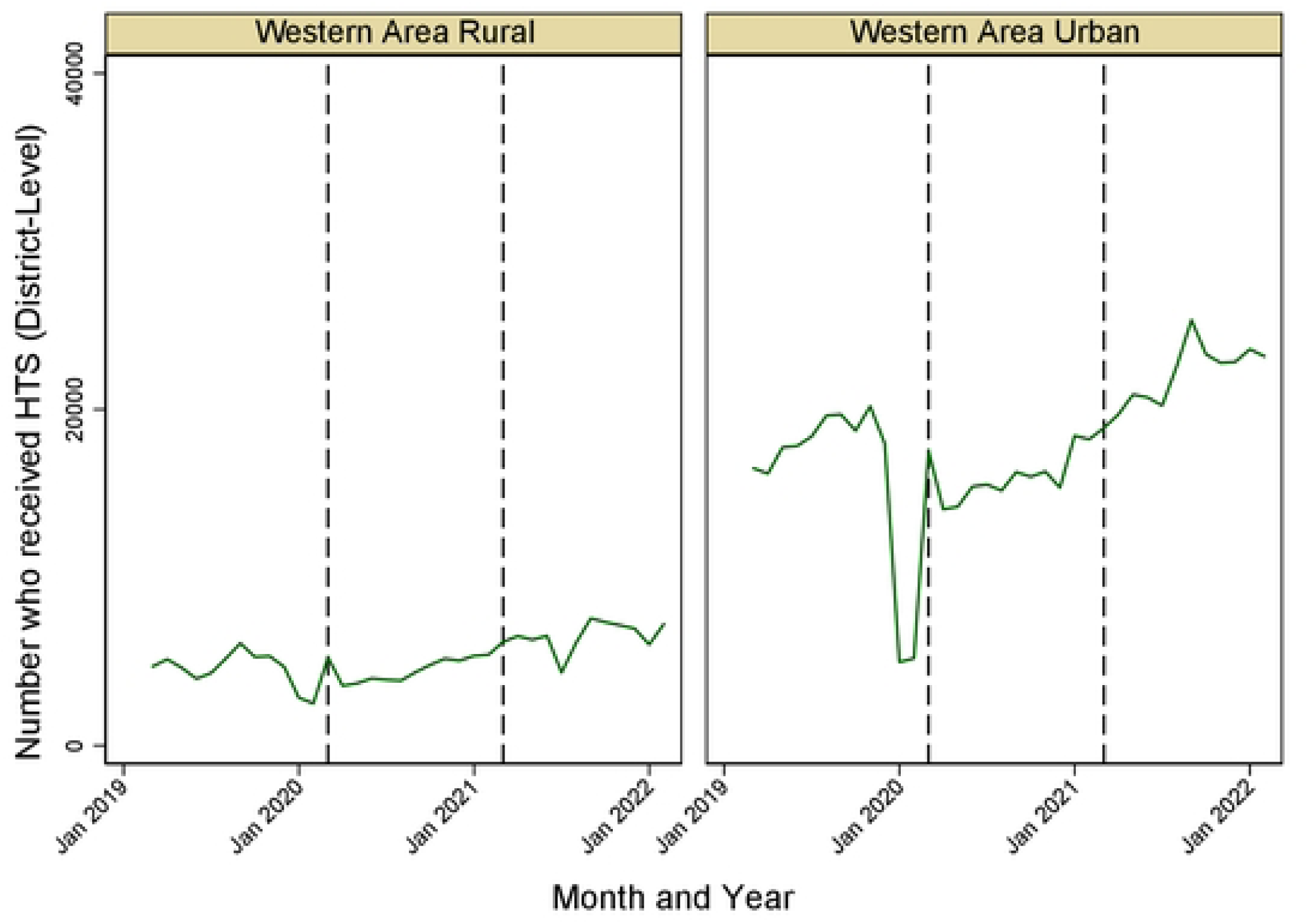
Variation in the utilization of HIV testing servicesacross Western Urban and Western Rural

## Discussion

We assessed the uptake of HIV testing services over a period ranging from pre COVID-19 pandemic to the post-COVID-19 era. Our results have revealed crucial patterns in the uptake HIV testing in the context of health system’s reactions to stress and adaptations. First, our results show an acute decline in the uptake of HIV testing services nationwide just before the first case of COVID-19 was reported in Sierra Leone. Second, urban areas were most affected, with Western Area Urban experiencing the most decline in HIV testing services uptake. Third, there was gender disparity in the utilization of HIV testing services. Finally, women use HIV testing services about twice as much as men but the decline in testing during the COVID-19 pandemic was similar for both sexes.

We found a decline in HIV testing uptake during the COVID-19 pandemic of 6.5%. Studies across low and middle-income countries have confirmed that the COVID-19 pandemic reduced HIV testing uptake among people living with HIV, but data to describe this effect in Sierra Leone is limited. In three facilities across the three different levels in the Western Sierra Leone, HIV testing services declined by 41% during the COVID-19 pandemic [9]. Our findings and the one reported in this formative study reflects on the global pattern of COVID-19 interruptions of health services. A study conducted by AIDS Health Care Foundation across four continents demonstrated a reduction in HIV service delivery across the continuum of care, including testing [10]. Research on the uptake of HIV testing services in Nigeria showed a significant decrease in uptake and linkage to care [11]. A study in India described a component of HIV testing services; index case testing revealed reduced service uptake and increased service cost [12].

Several factors could have explained the reasons for the decline in HIV testing services. One of the factors contributing to the acute decline in HIV testing services was the industrial action by health workers, which was reported during the COVID-19 pandemic as health workers demand for risk allowance and other compensations in response to the pandemic. This situation is not unique to Sierra Leone. In Kenya, a national strike by health workers resulted in significant reductions in essential services such as hospital deliveries and caesarean sections in public hospitals during the COVID-19 pandemic [13]. Therefore, we call for investment in the heath workforce as part of health system building strategy to prevent the disruptions in service delivery in future public health emergencies.

The temporary suspension of index case HIV testing to mitigate the spread of COVID-19 may have led to a decline in HIV testing services. Incorporating HIV testing into COVID-19 treatment centers may have helped offset this reduction, but due to the low number of COVID-19 cases in Sierra Leone, HIV testing services were still disrupted by the pandemic.

HIV testing recovered rapidly in the wake of the COVID-19 pandemic, which can be attributed to the adaptive measures taken during the pandemic. These observations suggest that health systems have the resilience to quickly adapt to shocks and restore essential services to pre-pandemic levels [14]. Experience with past epidemics, particularly the Ebola outbreak, and differences in population dynamics are critical considerations when developing emergency response plans [15]. Building a strong health system is important to reduce a country’s vulnerability to public health emergencies and ensures a high level of preparedness to mitigate the impact of any crises. Sierra Leone’s ability to rapidly bounce back from health system shock and maintain HIV services amidst multifaceted health systems challenges underscores the evolution of resilience in its healthcare system. Our findings highlight broader lessons for global health, underscoring the importance of integrated and resilient health systems that are able to manage multiple health threats simultaneously while ensuring equitable access to essential services for all populations, especially in times of crisis.

We report regional differences in service provision. HIV testing services in urban areas, especially in the Western Area Urban, have been more affected than in provincial or rural areas. This difference may explain why HIV testing services have changed little or not significantly in rural areas compared with urban areas. Community behavior and engagement strategies differed between urban and rural areas: urban areas used mainly electronic, print and social media for health promotion, while rural areas benefited from road shows and direct community engagement, with relatively high degrees of impact on the effectiveness of these approaches.

There were more stricter restrictions in the Western Area compared to other regions. Finally, in rural areas, the provision of health services depends not only on clinic visits but also on home services provided by community health workers [16].

Women are particularly vulnerable to the impact of the COVID-19 pandemic [17]. Although more women than men undergo HIV testing in our study, the impact of the epidemic on HIV testing did not differ.

Our findings have limitations inherent to operational research using secondary data. There is limited reporting on DHIS2 for private hospitals. Therefore, the findings reported in this study are not representative of the impact of COVID-19 on HIV testing services in private settings in Sierra Leone. Importantly, we used secondary data in DHIS2 and could not control the quality of data input into this database. The use aggregate data limited our ability to analyse relevant variables such as age groups and sub-populations. Finally, we have not been able to determine cause and effect relationships of the impact of COVID-19 on HIV testing services.

Despite these limitations, there are several strengths of this study. First, we analyzed data generated from all regions and districts in Sierra Leone to make our findings nationally representative. Second, because the data we used were collected during routine operations and were not influenced by any induced observations during the collection period, they provide a fair reflection of the performance of the Sierra Leone health system under stress. Finally, while most studies focus on only the pandemic period, our study extends the scope to a broader timeline, covering the pre-, during-, and post-COVID-19 pandemic phases. Thus, adding to the burden of evidence that calls for the protection of essential health services during public health emergencies.

**Conclusion:** Our findings highlight that HIV testing was significantly disrupted during the COVID-19 pandemic but recovered quickly after the pandemic. These findings highlight the lessons learned from previous epidemics and the importance of adaptive strategies to maintain essential health services during public health emergencies. Future research should explore the specific factors that influence health system adaptability and resilience in the context of a public health emergency to inform effective public health interventions and policies.

### Declaration section

The authors declared no conflict of interest

## Data Availability

The datasets (https://doi.org/10.5061/dryad.3r2280gsp) used and/or analyzed for this study are available from: http://datadryad.org/stash/share/A7iknh0neWpaUg4eUBL3fuO8Dcmgpjm31FFVZwtacf8

## Acknowledgements

SM is supported by a Tier 2 Canada Research Chair in Mathematical Modeling and Program Science.

## Author’s contribution

Conceptualization and study design: HS, GE, and SL

Data curation and validation: BM, FKT, JT

Methodology and formal analysis: HS, SM and GE

Funding acquisition, resources, supervision: SS, SL, SM, ACK

Writing – original draft preparation: HS, SS, ACK and GY

Writing – review & editing: SL, ACK and SM

**Availability of data:** The datasets (https://doi.org/10.5061/dryad.3r2280gsp) used and/or analyzed for this study are available from: http://datadryad.org/stash/share/A7iknh0neWpaUg4eUBL3fuO8Dcmgpjm31FFVZwtacf8.

